# The TIDIER Trial: A randomized, phase II clinical trial of time-restricted eating versus nutritional counseling among patients receiving radiation or chemoradiation for prostate, cervical or rectal cancer

**DOI:** 10.64898/2025.11.28.25340848

**Authors:** Ziyi Huang, Errol J. Philip, Jonathan Anzules, Leslie Wenning, Greisha L. Ortiz-Hernandez, Nathan Lin, Qianhua Feng, Chenying Ma, Paul Frankel, Tanya Dorff, Yun Rose Li

## Abstract

**Background:** Radiation therapy (RT) is an essential component of definitive treatment for cancers of the prostate, rectum, cervix, and other pelvic malignancies. Despite its proven efficacy, pelvic RT is associated with a constellation of adverse effects, collectively referred to as pelvic radiation disease (PRD). Several clinical and preclinical studies have shown that dietary interventions, including time-restricted eating (TRE), may represent a novel approach to mitigate radiation and chemotherapy toxicity and, in some cases, promote oncologic efficacy, thereby enhancing quality of life, reducing morbidity and mortality, and improving disease outcomes.

**Objectives:** This trial was designed to evaluate TRE feasibility/tolerability and test the hypothesis that TRE during RT or chemoradiation could reduce DNA damage accumulation in peripheral blood mononuclear cells over the course of treatment, as quantified by γH2AX foci. Secondary objectives include reduction in clinically observed toxicities, improvements in urinary or blood biomarkers of DNA damage, mitigation of microbiome dysbiosis and cytokine response during radiation, and improved metabolic parameters.

**Methods:** A total of 48 individuals with prostate, cervical, or rectal cancer aged ≥18 years with a BMI ≥21 kg/m^2^ receiving pelvic radiation will be randomized to either TRE or nutritional counseling. Both groups of participants will receive dietary consultation with a registered dietician before and during RT, and the TRE group participants will be asked to restrict their caloric intake to a specific timeframe (fast at least 6-8 hours before and at least 4-6 hours after RT). Biospecimens (blood, urine, stool), quality-of-life surveys, physician-reported toxicities (CTCAEv5.0), and food intake and fasting logs will be collected biweekly during the study intervention and every 3-6 months up to one year follow-up.

**Results:** Accrual was completed in August 2025. Follow-up and biospecimen collection are ongoing, and correlative analyses are in progress. Final outcomes will be reported upon study completion.

**Trial registration:** ClinicalTrials.gov NCT05722288

## Background

Radiation therapy (RT) is an essential component of the definitive treatment for advanced and high-risk cancers of the prostate, rectum, cervix, and other solid malignancies of the pelvis. Typically, pelvic RT consists of approximately 5 weeks of treatment (over 25-30 individual sessions) to a dose of 44-54Gy, with the intent of covering both the primary tumor and pelvic lymph node basins at risk of microscopic metastatic disease. Whereas concurrent systemic therapy differs, cancers of the prostate, rectum, and cervix are treated very similarly from a radiation standpoint. Despite its proven efficacy, pelvic RT is associated with both acute and late side effects, including gastrointestinal and genitourinary symptoms resulting from inflammatory damage, neovascularization, and fibrosis, as well as secondary malignancies [1, 2].

The constellation of adverse effects that can result from pelvic RT has been collectively referred to as pelvic radiation disease (PRD) [3]. PRD is more common than previously recognized; in some studies, 90% of patients who received pelvic RT reported a chronic change in their bowel habits, and 50% reported a significant change in their quality of life [4]. Clinically, PRD is a highly heterogeneous phenomenon, as many patients’ intrinsic factors may impact normal tissue response and experience of toxicity. These factors include differences in dietary intake (e.g. fiber intake, microbiome health), medical comorbidities that impact bowel health (e.g. inflammatory bowel disease), anatomy (bowel distribution), as well as variations in radiation treatment planning and delivery (e.g. use of daily IGRT, IMRT technique, extent of local disease) [3, 5].

Prior work has suggested that fasting may be associated with fewer treatment side effects in the setting of chemotherapy [6, 7]. Notably, fasting has been shown to reduce DNA damage in normal tissue in prior clinical trials of patients undergoing chemotherapy [8, 9]. In contrast, tumor cells, which do not have effective DNA repair machinery and are less able to adapt to low nutrient conditions, remain more vulnerable in the setting of fasting. As both the efficacy and toxicity of RT, similar to chemotherapeutic agents like cisplatin, are in large part due to oxidative cellular and DNA damage [10–12], this effect may thus improve the therapeutic ratio of treatment and represent fasting as a novel approach to both limit toxicity and promote treatment efficacy in the setting of cancer treatment.

The evidence noted above prompts the question of whether fasting could be beneficial for patients undergoing RT. Cell culture experiments, which have documented reduced radiation toxicity to healthy cells in a low-glucose medium, revealed that neoplastic cells do not possess the same innate protection as healthy cells and thus may be sensitized to RT-induced cytotoxicity in this environment [13, 14]. Pre-clinical studies involving mice have also revealed that short-term starvation (24 hours) or prolonged time-restricted eating (TRE) significantly reduced the toxicity of treatment and improved the mortality rate from high-dose RT compared to mice fed standard diets prior to RT treatment [15, 16]. Moreover, histologic studies of mice receiving abdominal RT showed that short-term fasting mice had higher numbers of crypts, increases in villi height, improved crypt regeneration, and higher levels of CC3 (a protein involved in DNA repair) expression compared to those fed normally [15]. However, there is limited data on the potential benefits of caloric restriction or TRE among humans receiving RT.

While TRE is well-tolerated in healthy adults, the tolerability of TRE is not well-documented in patients with cancer, particularly those receiving intensive daily therapy such as pelvic RT. The primary objective of this trial is to evaluate the feasibility and tolerability of TRE among patients receiving long-term daily radiotherapy for pelvic malignancies.

As a co-primary objective, we aim to test the hypothesis that TRE can reduce DNA damage during RT or chemoradiation therapy, as quantified by γH2AX foci. γH2AX is a sensitive marker for DNA double-strand breaks (DSBs), which could be induced by various factors such as ionizing radiation, reactive oxygen species (ROS), and deficient repair of other non-DSB DNA lesions [17–19]. Previous studies have demonstrated increased induction and persistence of γH2AX, measured by flow cytometry, in patients who experienced severe atypical normal tissue toxicity (NTT) compared to those with minimal or no NTT [20]. These findings support the potential of γH2AX as a predictive biomarker for treatment response and RT-related toxicities.

The impact of TRE on other biological substrates, including microbiome diversity and the development of radiation-induced microbiota dysbiosis, is a further area of emerging research. Prior preclinical and non-oncology clinical studies have shown that various forms of TRE may be associated with enhanced diversity and richness of microbiota [21, 22]. Meanwhile, pelvic radiation is associated with the depletion of gut microbiota diversity and the relative abundance of individual bacterial taxa [23]. These opposing effects highlight a critical and understudied gap in knowledge of whether TRE can mitigate the adverse effects of radiation on the gut microbiome, thereby reducing treatment-related gastrointestinal side effects. Despite a compelling biological rationale, prospective clinical data investigating the combined effects of TRE and pelvic RT on the microbiome remain limited. To address this gap, our study will longitudinally characterize microbiome changes in patients undergoing pelvic RT and evaluate the influence of TRE on microbial composition and diversity throughout treatment.

Androgen deprivation therapy and corticosteroids associated with chemotherapy administration could adversely impact patient metabolism. Meanwhile, fasting results in prompt metabolic adaptation. Studies in healthy volunteers demonstrated significant metabolic changes, including reductions in blood glucose and insulin, and increased ketone levels, within just 22-48 hours of fasting [24, 25]. Preclinical and clinical studies in mice and humans also demonstrated a reduction of insulin-like growth factor 1 (IGF-1) of up to 75% and changes in the metabolic dynamics of its binding proteins after short-term fasting (48-120 hours) [26–28]. Decreased activity of the IGF-1 signaling pathway has been shown to protect simple organisms and cells against a variety of toxins and is expected to promote resistance to oxidative stress, as supported by pre-clinical data [29]. Several clinical studies suggest that fasting can activate protective signaling pathways, thereby enhancing metabolic hemostasis and reducing comorbidities such as diabetes and cardiovascular diseases [30–32]. We anticipate that fasting glucose, insulin, and/or IGF-1 will change from the baseline levels in the enrolled subjects, and we will evaluate whether TRE can alleviate this effect.

The cellular response to fasting and RT is a multilayered and precisely coordinated biological process that remains incompletely understood. In addition to the previously mentioned mechanisms, RT has been shown to induce cancer cell death and autophagy, leading to the release of tumor-associated antigens and the enhancement of targeted immune response. Concurrently, RT also damages normal tissues through the generation of ROS, contributing to treatment-related toxicity [12]. These observations underscore the importance of incorporating metabolomic and proteomic analyses in this trial to comprehensively map the complex network of molecular signaling and systemic responses within the human body.

### Protocol (Methods and analysis)

1. Study design

This is a single-institution Phase II randomized clinical trial registered on the clinicaltrials.gov website (ID: NCT05722288). The trial is conducted at City of Hope Medical Center (Duarte, CA) under the direct supervision of the Institutional Review Board of the institution.

Patients enrolled must be diagnosed with localized prostate cancer, rectal cancer, or cervical cancer patients receiving pelvic radiation therapy (RT) or chemoradiation therapy (CRT) and intend to receive similar total biological doses of pelvic radiotherapy over a similar duration of treatment time, as well as similar anatomic treatment fields (45-54 Gy in 25-30 fractions to pelvic lymph nodes and disease site), and are expected to experience similar areas/domains of treatment toxicity. The study flowchart of enrollment, allocation, intervention, and follow-up is presented in Figure 1.

**Figure 1.**
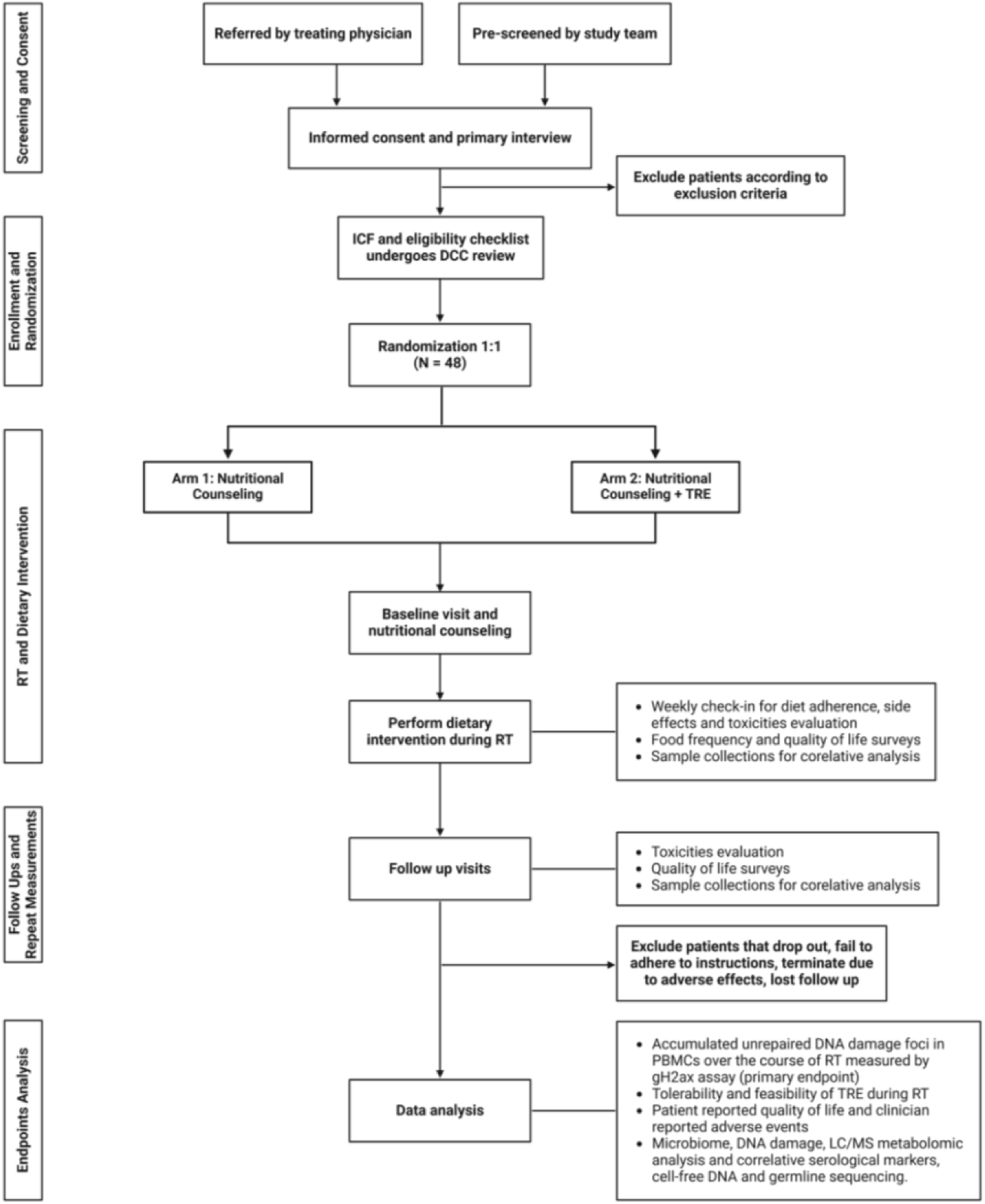
Study Workflow. Patients with prostate, rectal or cervical cancer will be referred from their treating physician or pre-screened by the study team. Signed consent forms and eligibility checklists will be sent to DCC for enrolling and randomization. Participants will be notified about their randomization result on the day of baseline visit and will perform dietary interventions only during their pelvic RT. **DCC**, Data Coordinating Center

Patients will be assigned to one of the 2 arms in a 1:1 fashion, with TRE or standard scheduled diet stratified by disease site. Dietary consultation and sample collection will be performed 1-2 weeks before starting RT or CRT during a baseline evaluation visit. Dietary interventions will be conducted every day the patient receives RT, and participants will be followed every 3 months until 1 year after finishing RT or CRT. A study calendar is presented in Figure 2.

**Figure 2.**
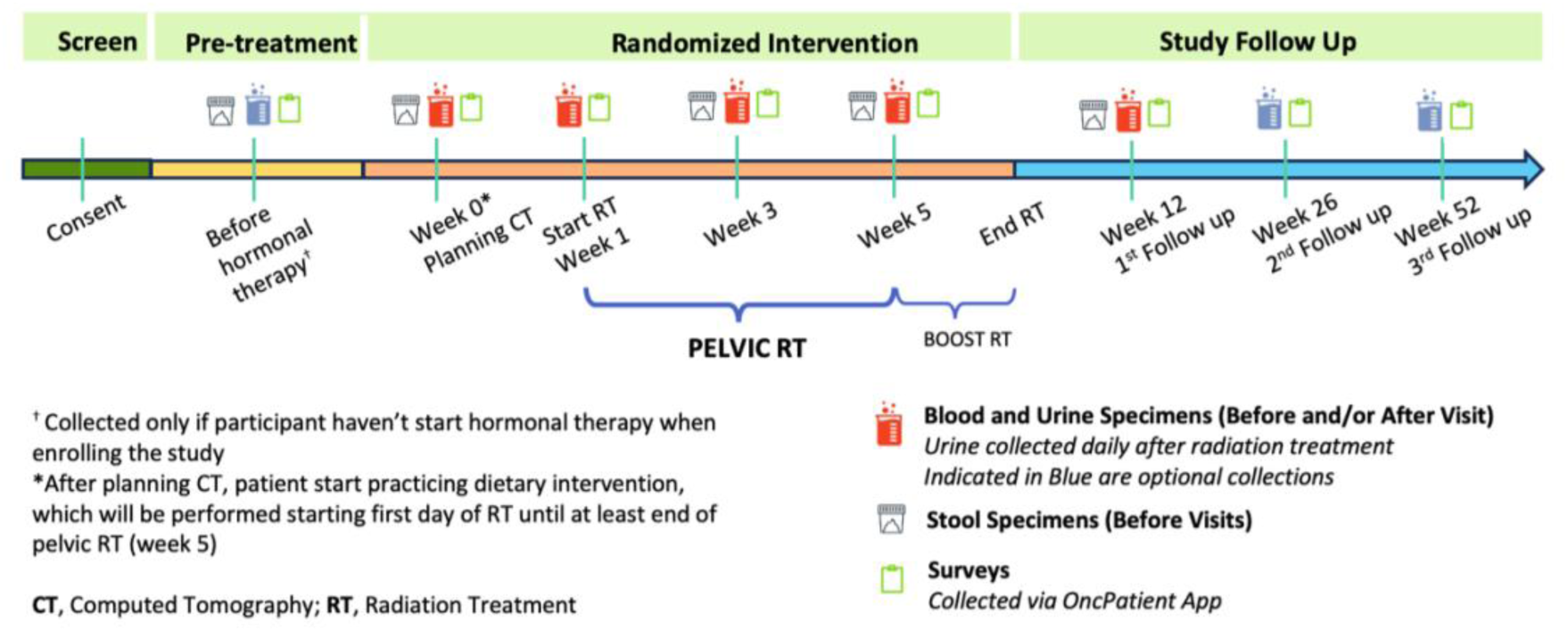
Study calendar.

**Figure 3.**
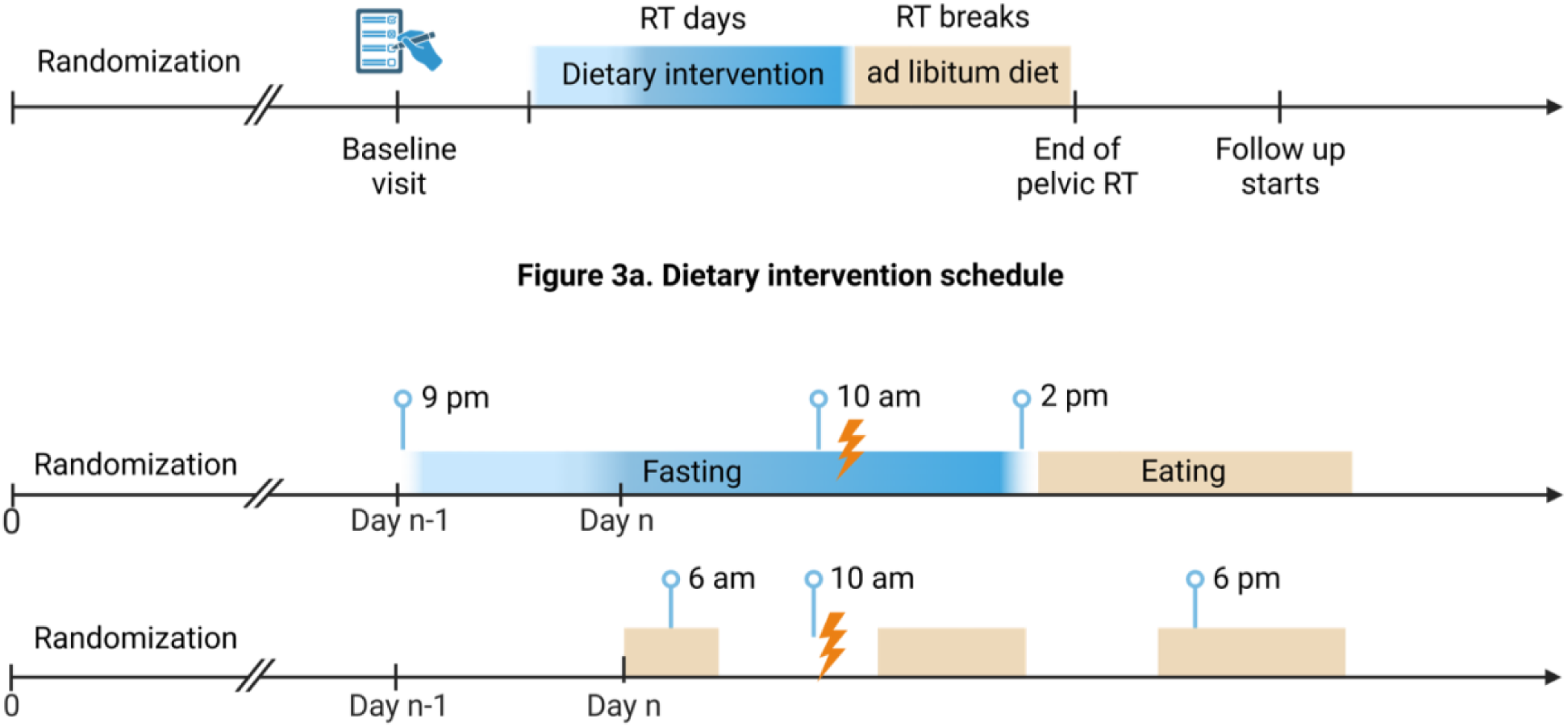
Dietary Intervention Regimen. 3a. Participants will perform dietary intervention only on days when receiving pelvic radiation. 3b. Comparing daily schedule of TRE group (above) with nutritional counseling group (below).

2. Sample size

The planned study recruitment will be forty-eight patients (N=24 per arm) with high-risk prostate cancer (stratum 1); local-regionally recurrent prostate cancer (stratum 2); cervical cancer (stratum 3); or locally advanced rectal cancer (stratum 4) undergoing radiation or chemoradiation therapy. Patients are expected to be accrued over the course of 36 months.

The primary study endpoint is the cumulative DNA damage of circulating peripheral blood mononuclear cells (PBMCs), as measured by the γH2AX assay over the course of RT (baseline compared to completion of pelvic RT at 5 weeks). Whereas there is limited clinical data on this assay, early pre-clinical data suggest that PBMCs are sensitive to radiation damage. Prior study sampling suggests that healthy controls have a positive γH2AX assay of ∼8%, while cells undergoing radiation have a several fold increase. We hypothesize that the accumulation of DNA damage will be lower among fasting patients. If the difference in the γH2AX assay is 5% (e.g., elevated from 8-10% in fasting and from 8-15% in non-fasting) and the standard deviation of this change is 7.5% (corresponding to an effect size of 0.667), a two-group t-test with a 0.1 one-sided significance level will have 84% power to detect this difference between the arms, with 24 patients per arm, for a total of 48 patients. To avoid imbalance between arms, patients will be stratified by disease: prostate (intact), prostate (post-prostatectomy), cervical, and rectal cancers. The actual analysis will use linear models that include the baseline and final γH2AX assay and stratification.

3. Participants Enrollment and Randomization

Study candidates will be pre-screened and referred by their treating physicians to the study team. After a consent form is signed and eligibility is confirmed, the enrollment package will be sent to City of Hope Data Coordinating Center (DCC) to review for enrollment and randomization.

4. Inclusion and exclusion criteria

For prostate cancer patients, only high risk or post-operative patients undergoing whole pelvic RT will be included. For cervical cancer, eligible patients will be those planning to receive definitive chemoradiation with FIGO Stage IB2 to IIIC; and for rectal cancer patients, only those receiving “long course” chemoradiation therapy over the course of 5 weeks will be included to avoid confounding introduced by use of different treatment fields and time courses. Inclusion and exclusion criteria are listed in Table 1.

**Table 1.** Eligibility criteria.

|  |
| --- |
| **<u>Inclusion Criteria:</u>** |
| Male and female patients aged 18 or older |
| Patients undergoing radiation or chemoradiation therapy with:<br>NCCN high risk or node positive prostate cancer;<br>Local-regionally recurrent prostate cancer after prostatectomy;<br>Locally advanced cervical cancer (FIGO stage IIb+); or<br>Locally advanced rectal cancer (cT3/4 or N+). |
| ECOG performance status 0-2 |
| Able to provide a written consent for study participation |
| **<u>Exclusion Criteria:</u>** |
| Body mass index (BMI) <21 |
| Prior pelvic radiation treatment |
| Women during pregnancy, lactation, or plan to become pregnant during study intervention |
| Diagnosis of insulin-dependent diabetic mellitus or poorly controlled diabetes |
| Currently engaging in strict macronutrient/time-limited diet (including ketogenic diets) |
| Unable to consent or unable to undergo time-restricted diet |

5. Intervention

Participants in both arms will receive pre-treatment dietary counseling with a dedicated dietitian and general guidance on a balanced macronutrient diet as calculated by their basal metabolic rate (BMR). Recommendations will be given to follow the American Institute for Cancer Research (AICR) healthy eating guidelines and adhere to a diet avoiding gas-promoting foods during radiation.

On days of RT (Monday through Friday), patients on the TRE arm will be asked to avoid caloric intake for a 12-14-hour duration beginning at least 6-8 hours before and ending at least 4-6 hours after RT; patients will be allowed an ad libitum diet on weekends (i.e. days they receive no RT). Patients in the control arm are not instructed to alter the timing of their caloric intake.

Water intake will not be limited during fasting. Participants will be advised to keep hydrated with water, coffee, or tea without sweeteners; broth (< 5 calories); or sugar-free drinks sweetened with natural sweeteners like stevia or monk fruit. When fatigue, weakness, hunger, headache, nausea, or other fasting-related discomfort limits their normal activities, participants will be advised to consume rescue foods no more than 1-2 times per fasting day to help them maintain the fasting period. Rescue foods are dietitian-approved items without added sugar that are limited to 50 calories.

Nutrition education information will be accessible on the Oncpatient Companion mobile application, and participants can revisit dietary recommendations whenever needed. The application also provides reminders to patients, collects food intake and fasting log data, and elicits patient feedback regarding tolerability.

6. DNA damage measurement

The primary goal of this study is to evaluate whether patients in the intervention arm possess reduced normal tissue DNA damage in the context of a TRE diet during RT. DNA damage will be measured by quantification of γH2AX foci, a type of persistent DNA damage that occurs after RT, using flow cytometry.

In addition to PBMCs, we will also examine if TRE reduces DNA damage observed in blood and urine based on the COMET assay for dsDNA breaks and ROS-associated DNA adducts as measured by 8-oxo-guanine mass spectrometry.

We will compare changes in γH2AX foci formation and concentration of 8-oxo-guanine adducts via an ROS DNA damage assay across serial time points for patients in the control and experimental groups using repeated measures. Measurements used for all assays will be an average based on triplicate measurements.

7. Feasibility and tolerability measurement

As a co-primary endpoint, feasibility/tolerability will be met if 70% of patients are able to compliant with the dietary intervention for 70% of the pelvic radiation treatment duration. Rescue food was allowed during the fasting period and will not considered an instance of noncompliance. Participants will log their fasting time and rescue food use in the Oncpatient Companion application. The study team will contact participants weekly to assess compliance and answer any participant questions.

8. Toxicities measurement

Toxicities during treatment and follow-up appointments will be measured by clinician-reported adverse events using CTCAEv5.0 and documented in a patient electronic health record system. Patient self-reported quality of life is collected by EORTC-PR25 (prostate cancer), EORTC-CX24 (cervical cancer), and EORTC-CR29 (rectal cancer) surveys through the Oncpatient Companion phone application and then documented in RedCap.

9. Metabolic changes measurement

Metabolic changes in both groups will be measured as the change in HbA1c, which we will compare at study baseline and at 12-week follow-up. Plasma will undergo metabolomics analysis to investigate and compare the metabolite changes during androgen deprivation therapy, RT, and follow-up.

10. Microbiome analysis

Microbiome of the gut will be analyzed for changes between baseline and 3 weeks of dietary intervention, as well as during the final week of RT and at a 3-month follow-up. Microbial diversity (Shannon Index) will be compared across all timepoints. Specific species associated with radiation dysbiosis will be evaluated for change using repeated measures ANOVA. These species include Lactobacillaceae, Lachnospiraceae, Ruminococcaceae, and Clostridiaceae [23].

### Study progress and implementation

To date, 51 eligible patients have consented to enroll in the study. Three patients withdrew before starting RT due to non-study-related side effects, such as emotional distress due to the overall treatment. Two patients were unable to provide biological samples or survey data and were replaced after starting treatment. The accrual is completed with forty-eight patients who have finished treatment and dietary interventions. Follow-ups and biospecimen collection are ongoing and estimated to be completed in August 2026. Final outcomes will be reported upon study completion.

## Discussion

Many forms of TRE interventions exist, including but not limited to daily 16:8 fasting, weekly 5:2 intermittent fasting, alternative day fasting, and the warrior diet. In designing this study, we considered several factors, including tolerability and biological rationale. We hypothesized that TRE around the time of RT could effectively enable normal stem cells to reduce metabolic sources of oxidative stress, which contribute to the overall free radical pool, thereby shifting cellular effort into DNA repair capacity in response to RT-related DNA damage. Because DNA repair is time-dependent, limiting the endogenous DNA damage induced by metabolic activity allows normal cells to avoid the accumulation of DNA damage, which would ultimately result in mitotic catastrophe. This, in turn, reduces treatment-related toxicity. Historically, when radiation is delivered twice or thrice a day, typical treatment includes a normal tissue recovery period of at least 4-6 hours. Thus, we stipulated that a fasting period consists of at least 6-8 hours before and 4-6 hours after RT, encompassing a total of 10-14 hours and including the typical 4-6-hour recovery window. The total duration is generally reported to be well-tolerated in most patients.

In addition, the hypothesis that prolonged nightly fasting could reduce cancer risk and improve cancer outcome was investigated in a prospective study with early-stage, invasive breast cancer patients enrolled in a large multisite randomized trial. This study demonstrated that a longer nightly fasting regimen was associated with significant improvements in glycemic control biomarkers and reduced both breast cancer recurrence and inflammation biomarkers [33–35]. While the underlying mechanisms by which such associations may exist are yet to be fully elucidated, study participants in the TRE group were encouraged to perform an overnight fast to further boost the potential benefits of TRE.

As studies have shown, the circadian rhythm plays a critical role in nutrition and homeostasis, and eating times, which are linked to the circadian rhythm, influence metabolic and inflammatory responses [35–37]. While commonly associated with nutrient availability, cell cycle repair is also intimately tied to individual circadian rhythms [38]. An emerging body of research suggests that the time of day at which therapy is administered may influence both treatment efficacy and toxicity [39–41]. Several studies, including those on lung, head and neck, colorectal, and breast cancers, have suggested that specific timing of both chemotherapy and radiotherapy may impact the toxic effects of treatment [39, 42, 43]. The potential interaction between fasting schedules and circadian rhythms may serve as a valuable topic of interest for future research to determine the effect of dietary timing on treatment outcomes. Based on the established critical role of circadian rhythm, all participants in this study will receive daily RT within a restricted timeframe (8am to 2pm). Consequently, patients who are unable to attend morning radiation sessions will be excluded.

Selection bias may be present in this study. Patients treated in tertiary centers may differ from the general population in socioeconomic characteristics and health awareness. Furthermore, individuals who choose to participate in a diet-related trial may have greater acceptance of fasting and, thus, higher adherence to the fasting regimen. These factors may contribute to inflated feasibility and tolerability outcomes as well as potential impact on reporting of radiation toxicities. All subjects will be encouraged to report all symptoms honestly and comprehensively to reduce this risk.

Another potential source of bias relates to the use of GLP-1 receptor agonists. Since their introduction, these agents have become an important aspect of diabetes management and weight loss therapy, demonstrating significant efficacy in treating metabolic disorders and providing protective effects for the cardiovascular system and kidneys [31]. A limited number of patients receiving GLP-1 receptor agonists were enrolled in this study. Given their mechanism of action, which includes a delay in gastric emptying and a reduced appetite, these medications may facilitate higher adherence to a TRE regimen. Based on the insight from this study, a follow-up randomized trial has been designed to compare the metabolic effect of TRE and GLP-1 receptor agonists. As part of the current trial, we will also collect data on potential correlates of TRE intervention in the setting of pelvic RT. Given the preliminary nature of fasting science in oncology, this data will provide insight into the potentially widespread and complex effects of TRE on treatment toxicity, psychosocial outcomes, metabolomic markers, and the gut microbiome.

γH2AX foci are well-characterized markers of persistent DNA double-strand breaks and are commonly used to assess DNA damage following RT. In this trial, quantification of γH2AX foci in PBMCs will serve as a co-primary endpoint to evaluate DNA damage and repair dynamics in normal tissue during treatment. This approach may offer mechanistic insight into inter-individual variability in radiation sensitivity and the development of acute and late toxicities. Notably, some of the most severe complications of RT do not occur until decades after treatment, thus complicating timely diagnosis and intervention. As the long-term survival of prostate and rectal cancer continue to improve, identifying patients at risk through non-invasive biomarkers like γH2AX, along with delineating approaches to mitigate risk in such patients, are essential to reducing PRD morbidity and mortality [44].

While prior studies outside of oncology have suggested that various forms of TRE are associated with enhanced microbiome diversity and richness, the impact of TRE on microbiome dynamics in the context of RT remains only partially elucidated [21, 22]. The longitudinal microbiome data collected in this trial will allow us to examine whether TRE can counteract radiation-induced microbial disruption and whether the preservation or restoration of microbial diversity correlates with reduced toxicity.

Furthermore, this study provides a unique opportunity to explore the microbiome as both a therapeutic target and a predictive biomarker. Such insights could be crucial for informing the development of microbiome-targeted strategies, such as dietary intervention, microbial therapeutics, and microbiome testing, to both predict and reduce PRD, and at the same time improve patient quality of life with an integrative approach.

Finally, the metabolic alterations and correlative serological markers examined in the trial, along with metabolomic and proteomic analysis, will further contribute to the identification of fasting-associated metabolic signatures, which may be associated with improved clinical outcomes. Such integrative multi-omics analyses will provide information on pathway-level changes associated with enhanced resilience against RT-induced toxicity.

In conclusion, this study tests the hypothesis that TRE during daily RT is a practical method that can reduce cumulative DNA damage measured by γH2AX foci in PBMCs. If successful, this trial will provide valuable insight about the role of dietary intervention in integrative cancer management and potentially improve patient quality of life during and following pelvic RT and CRT. At the same time, this work will help establish the foundations for novel therapeutic combinations and identify new biomarkers to assess and predict treatment outcomes in cancer patients.

## Data Availability

All data produced in the present study are available upon reasonable request to the authors

